# Investigating idiopathic anosmia as a prodromal state of Parkinson’s disease

**DOI:** 10.64898/2025.12.17.25342478

**Authors:** Richard Nathaniel Rees, Sophie I Meyer, Carl Philpott, Simon Gane, Lisha McClelland, Cristina Simonet, Jonathan P Bestwick, Alastair J Noyce, Anette Schrag

**Affiliations:** Department of Clinical & Movement Neurosciences, UCL Queen Square Institute of Neurology, University College London, London, UK; Chelsea & Westminster Hospital NHS Trust, Imperial College London, London, UK; Centre for Preventive Neurology, Wolfson Institute of Population Health, Queen Mary University of London, London, UK; Norwich Medical School, UEA, Norwich, UK; Norfolk Smell & Taste Clinic, Norfolk & Waveney ENT Service, Norfolk, UK; Royal National ENT and Eastman Dental Hospitals, University College Hospital, London, UK; ENT, University Hospitals Birmingham NHS Foundation Trust; Homerton Healthcare NHS Foundation Trust, London, UK; Department of Neurology, The Royal London Hospital, Barts Health NHS Trust, London, UK

## Abstract

**Background:** Loss of smell commonly predates the diagnosis of Parkinson’s disease (PD). However, smell loss has multiple causes, and the relationship between idiopathic anosmia (IA) and PD remains incompletely understood.

**Objectives:** To assess the presence of prodromal features of PD in individuals with IA and to examine the relationship between anosmia duration and prodromal features.

**Methods:** Within the PREDICT-PD study, patients with IA investigated at specialist smell clinics were compared with healthy controls at low risk of PD (HC) and patients with PD. In-person assessments included MDS-UPDRS I-III, functional motor tasks, cognitive tasks, autonomic symptoms, orthostatic hypotension, pain, sleep and mood. We compared these features and PD risk according to the PREDICT-PD algorithm between the groups and examined the relationship between the duration of anosmia and severity of prodromal features using linear regression models.

**Results:** We recruited 42 IA, 28 HC and 22 PD participants, matched for age and gender. Overall motor scores were not different between IA and HC. IA had worse cognition, sleep, pain, depressive and some symptoms of dysautonomia than HC, with trends towards worse anxiety scores and higher PD risk scores. Shorter duration of anosmia was negatively correlated with MDS-UPDRS II scores but no other prodromal feature.

**Conclusions:** Some individuals with IA exhibit non-motor features suggestive of prodromal PD. This supports the known association of olfactory dysfunction with subsequent PD but suggests that, even after excluding secondary causes of anosmia, other conditions may underlie anosmia or smell loss may be temporally remote from PD.

## Introduction

The link between olfactory dysfunction and Parkinson’s (PD) was established nearly 50 years ago.^1^ However, hyposmia is common in the general ageing population: 12.4% of older adults in the US reported reduced smell and 22% had objective olfactory dysfunction.^2^ Typically, clinical history, examination and further investigations reveal the underlying cause of hyposmia. Idiopathic anosmia (IA) is diagnosed when acquired smell loss has no clear precipitant (like sinonasal infection or trauma) and is not linked to systemic disease.^3–5^ In an audit of diagnoses from a UK smell clinic with over 400 patients, chronic rhinosinusitis was most commonly diagnosed (40%), with idiopathic and post-viral dysfunction also common (30% and 17% respectively).^6^

Hyposmia is present in around 70% of PD patients at diagnosis, and up to 90% during the disease course.^7,8^ In the Honolulu-Asia Ageing Study, 2,267 neurologically healthy elderly men were smell tested at baseline and followed for 8 years. There was a five-fold increased risk of PD conversion in the bottom olfaction quartile compared to normosmic participants in the first 4 years, but this association was lost after that.^9^ Three large population-based prodromal PD studies have also used olfactory dysfunction as a risk marker: the Tübingen Evaluation of Risk Factors for Early Detection of Neurodegeneration (TREND) study,^10^ Prospective Validation of Risk Factors for the Development of Parkinson Syndromes (PRIPS)^11^ and Parkinson At Risk Study (PARS).^12^ The largest, PARS, used postal smell testing to invite participants for clinical follow-up. Minimal ascertainment of the cause of smell loss meant common causes were not rigorously excluded. Hyposmia is included as a risk marker in the International Parkinson’s Disease and Movement Disorders Society (MDS) research criteria for prodromal PD diagnosis,^13,14^ and forms part of the ‘prodromal’ recruitment drive for the Parkinson’s Progression Markers Initiative (PPMI).^15^

Both PPMI and PARS identified associations between olfactory loss and α-Synuclein seed aggregation assay (α-syn SAA) positivity.^15,16^ PARS reported α-syn SAA positivity in 48% (34/71) of hyposmic participants, compared to 4% (1/25) of normosmic individuals, while PPMI detected α-syn in 88% (16/18) of their hyposmic group. A subsequent clinic-based study further demonstrated co-occurrence of SAA positivity with both IA and prodromal Lewy body features.^17^ These associations suggest anosmia may not just be a clinical feature of PD, but the result of early synuclein disease processes. The use of α-Synuclein as a biomarker supports the biological definition of PD as a synucleinopathy and may aid disease-staging efforts.^18^ It may also aid IA patient stratification before motor symptoms occur, and account for the heterogeneity seen in this group.

Based on an assumption that systemic, genetic, iatrogenic and other causes of anosmia are unlikely to be PD related, we assessed the relationship between *idiopathic* anosmia and PD risk. We recruited patients with total or functional anosmia, assessed in specialist ENT-led smell clinics, to the PREDICT-PD study. This is a UK-based longitudinal study of older adults, remotely assessing known risk and prodromal features with a validated algorithm to determine individual PD.^19,20^

We hypothesised that individuals with IA would have more motor impairment and an increased frequency of non-motor features of prodromal PD than healthy controls (HC). We further hypothesised that IA prodromal features would be inversely correlated with duration of smell loss, suggesting developing neurodegeneration, rather than long-standing, gradual-onset, non-neurodegenerative causes of anosmia.

## Methods

All participants gave informed written consent, and the study received ethical approval from the London Queen Square (13-LO-1457, 10/H0716/85) and London Bloomsbury (17-LO-2021) Research Ethics Committees.

### Setting

IA participants were recruited from three UK sub-specialist otorhinolaryngology smell and taste clinics having had psychophysical smell testing, expert clinical assessment and relevant investigations concluding negative/normal for other causes of hyposmia (including imaging and endoscopy). PD participants were recruited through NHS movement disorders clinics in South-East England. HC were recruited through the PREDICT-PD study, which is described elsewhere.^19,20^ Through PREDICT-PD, all participants completed online lifestyle, medical history and non-motor symptoms questionnaires.

### Participants

Inclusion criteria:

1. IA - age >55 years, no identifiable aetiology on history or examination, negative nasal endoscopy, no significant pathology seen on CT or MRI scan (other than reduced olfactory bulb volume), absence of positive findings on blood screening for rare metabolic causes.
2. PD - idiopathic PD diagnosis from a consultant neurologist using UK Brain Bank criteria.^21^
3. HC - age ≥60 years, identified as being at low risk of PD (range of PREDICT-PD study algorithm estimated odds of PD: 1:60 - 1:30218, with a median of 1:2243).

Exclusion criteria:

1. IA - improvement of olfaction to oral corticosteroids, Sniffin’ Sticks TDI score of >31, evidence of concomitant sinonasal pathology.
2. PD - other cause of Parkinsonism, other neurodegenerative disease.
3. HC - presence of neurodegenerative or sinonasal disease.

### Data sources/measurement

Anosmia patients were assessed as being anosmic or functionally hyposmic in a specialist smell clinic – using a combination of patient history and psychophysical testing. These tests were performed according to clinical need and availability. All participants completed the PREDICT-PD online questionnaires. This allowed the calculation of PD risk, expressed as an odds, based on age and risk and protective factors with positive and negative likelihood ratios derived from meta-analyses of the literature.^22^ Olfactory function was excluded from the PREDICT-PD risk algorithm for the present study. Both participants and assessor were blinded to their risk profile. This algorithm has been published elsewhere and tested in other populations.^19,20^

All participants were assessed with the MDS-Unified Parkinson’s Disease Rating Scales (MDS-UPDRS) I-III. Functional motor assessments included the timed 3m Up & Go test (TUG), a test that changes with PD progression and discriminates between PD and healthy older controls.^23^ Subthreshold parkinsonism was defined as MDS-UPDRS III score >6, not including postural or action tremor.^13^ To assess handwriting, as a test of fine motor control, bradykinesia and higher cognitive functioning,^24^ participants copied the sentence: “Mary had a little lamb, its fleece was white as snow” three times. Each iteration was timed, starting with the initial pen-stroke and finishing at the end of the final word. The sentences were measured in cm (to the nearest 1mm), from the left-hand most mark to the right-hand most mark. Writing speed was calculated as the average length (cm) ÷ average time (s).

Cognitive tests included the Montreal Cognitive Assessment (MoCA), verbal fluency (letters F, A and S) and semantic fluency (animals) (with raw scores converted to age- and education-adjusted z-scores using published normative data), as well as the symbol digits modalities test (SDMT).^25–27^ Due to a change in the protocol to include verbal and semantic fluency tests, and the SDMT, in the HC group only 11, 11 and 24 (out of 28) completed these tests respectively, with 18, 24, 24 (out of 42) IA, and 15, 18 and 21 (out of 22) in the PD group.

Autonomic tests included SCOPA-AUT^28,29^ and measurement of supine and erect blood pressure at 1 and 3 minutes after 2 minutes supine rest. Orthostatic hypotension was defined as a drop of ≥20mmHg systolic or ≥10mmHg diastolic. Sleep was assessed with the Parkinson’s Disease Sleep Scale (PDSS),^30^ with the REM Sleep Behaviour Disorder Questionnaire (RBDSQ) forming part of the online battery.^31^ For pain, three questions from the King’s Parkinson’s Pain Scale (KPPS) were chosen for their discriminatory value in the clinometric analysis of the questionnaire.^32^ These were joint pain, pain turning in bed, and neuropathic or radicular pain. Mood was assessed with the Hospital Anxiety and Depression Scale (HADS).^33^

### Statistical methods

The IA group was compared to the HC and PD groups using two-sided t-tests or Wilcoxon rank sum tests for non-parametric variables. Shapiro-Wilk’s and Levene’s tests assessed normality and homogeneity of variance. Significance was set at p<0.05 and, unless otherwise stated, p-values were reported uncorrected. For tests of proportions, chi-square tests were used. Associations between anosmia duration and prodromal features were assessed using univariate linear regression and age- and sex-adjusted multivariate models. Data processing and analyses were completed using R (version 3.6.2).^34^ This study is reported following STROBE guidelines for observational studies.

## Results

### Participants

The study included 92 participants: 42 IA, 28 HC, and 22 PD. A further 26 individuals were contacted. Of these, one died after referral, three declined to be involved, 18 did not respond and four visits were cancelled due to the COVID-19 pandemic. IA participants were assessed in their own homes. All HC (n=28) and PD (n=22) were seen at the National Hospital for Neurology and Neurosurgery. PD participants on dopamine replacement therapy were examined in the ON medication state. Due to the timing of the data lock, 12 IA and 10 PD participants did not have complete HADS data.

### Descriptive data

The groups were well matched for age and gender (table 1). The IA group had a lower mean education level than the HC group (14.6 vs 17.5, p=0.009), although on average all groups had completed secondary education. Duration of smell loss in the IA group ranged from a few months to over 5 decades (median 7 years, IQR 8). In the PD group, the mean disease duration ranged from 1 to 15 years (median 6, IQR 8). Further baseline characteristics are summarised in table 1.

**Table 1.** Baseline characteristics of cohort. HC: healthy control, IA: idiopathic anosmia, PD: Parkinson’s disease, LEDD: Levodopa equivalent daily dose. Data are mean (SD)*, median (IQR) † or percentage (number). Two-sided t-tests were used for parametric data, Wilcoxon rank sum tests were used for non-parametric data and chi-squared tests for tests of proportions.

| Group<br>n | HC<br>28 | IA<br>42 | PD<br>22 | *p*<br>- |
| --- | --- | --- | --- | --- |
| Age (yrs)* | 72.3 (5.6) | 70.1 (5.4) | 72.0 (5.1) | 0.280 |
| Gender (male) | 50% (14) | 52.4% (22) | 59.1% (13) | 0.805 |
| Education (yrs)* | 17.5 (4.0) | 14.6 (3.2) | 16.1 (4.2) | 0.007 |
| Disease Duration (yrs) † | - | 7 (8) | 6 (8) | - |
| LEDD (mg)* | - | - | 503 (416) | - |

### Main results

#### Comparison of the IA group to the HC and PD groups

The IA group did not differ significantly from HC, with similar MDS-UPDRS III scores, writing speed and the TUG. As expected, the PD group was significantly more impaired than the other groups (figure 1, supplementary table 2). 10/42 (23.8%) IA had subthreshold parkinsonism, which was not significantly different to controls (3/27 (11.1%), p=0.224). MoCA scores were lower in the IA group compared to HC (25.5 vs 27, p=0.019) but not different to the PD group. There were no significant differences between IA and either HC or PD in the extended phonemic fluency tests or the SDMT, although the IA group had worse performance than PD but not controls on the expanded verbal fluency (−0.04 vs 0.83, p=0.012).

**Figure 1.**
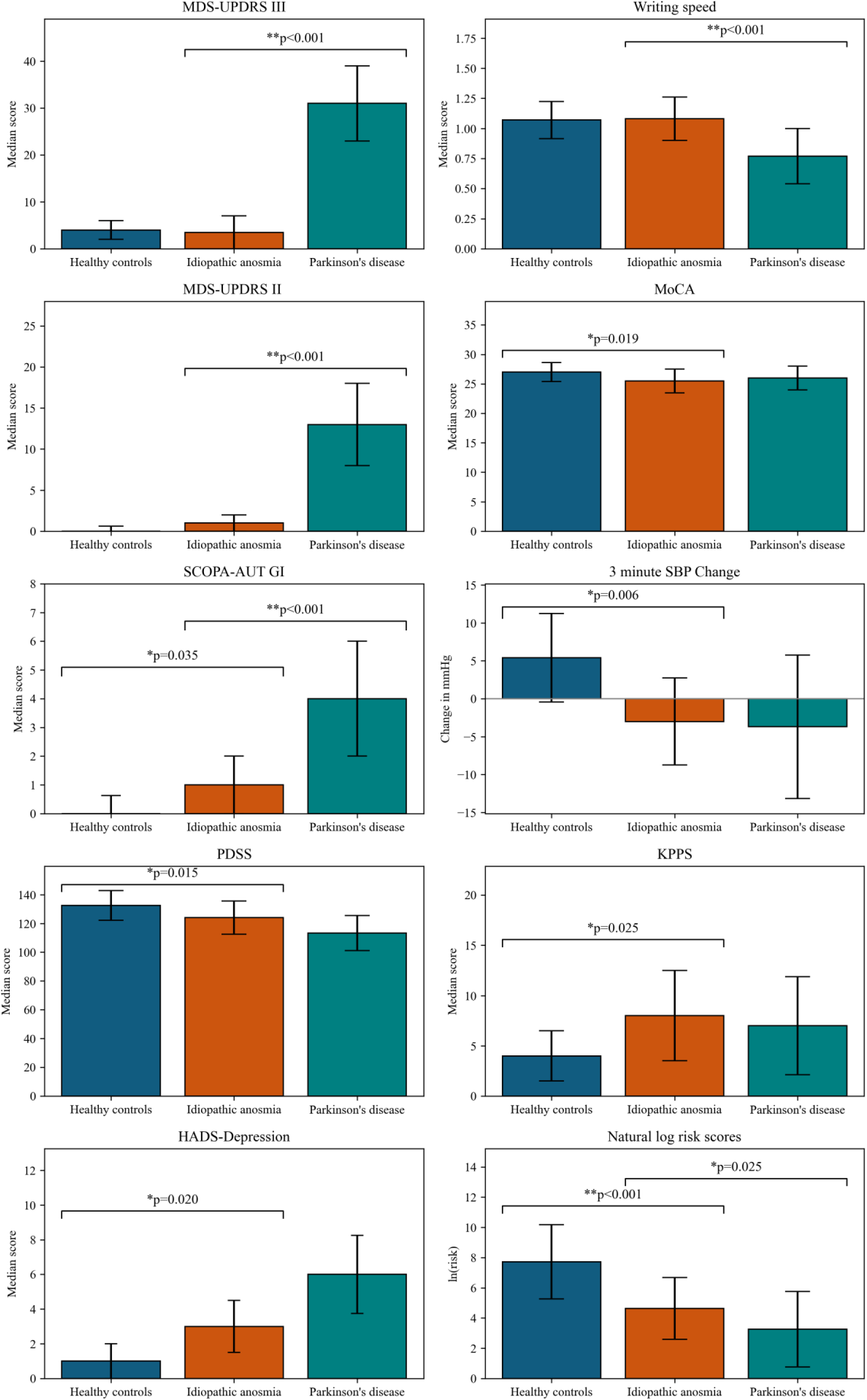
Median (IQRs), mean (SD) and natural log risk scores across groups. For variable-level group n, please see supplementary material table 3. Statistically significant p-values (p<0.05 *, p<0.001 **) derived from Wilcoxon rank sum tests.

There were no differences in autonomic symptoms in the total SCOPA-AUT between HC and IA, but there was a higher score in the PD group than IA (13.2 vs 9.08, p=0.021) (supplementary table 2). However, in the GI subdomain, IA had a small but significant increased symptom burden than HC (1 vs 0, p=0.035, figure 1), and PD were higher than IA (4 vs 1, p<0.001). While no group showed orthostatic hypotension at 1 or 3 minutes, the IA group had a greater systolic drop than HC at 3 minutes (−3.0 vs 5.4, p=0.006, figure 1), and PD were slightly more affected than IA at one minute (−11.9 vs −2.7, p=0.035) with no difference at 3 minutes.

PDSS scores revealed more sleep dysfunction in the IA group, compared to the HC group (124.1 vs 132.5, p=0.015), with a trend towards worse scores in the PD group (p=0.084) - lower scores indicating worse sleep (figure 1). There were also higher IA pain scores compared to the HC group (8 vs 4, p=0.025), with no difference from the PD group.

Few participants had scores outside the normal range for anxiety or depression on HADS (supplementary table 1). However, the IA participants had higher depression scores than HC (3 vs 1, p=0.02, figure 1) with a trend towards higher scores in the PD group, and a trend towards higher anxiety scores in the IA group than the HC group with no difference to the PD group (figure 1).

MDS-UPDRS I and II scores did not differ between the IA and HC groups, with higher scores in the PD group than the IA group (I: 11.71 vs 7.29, p=0.020, II: 13 vs 1, p<0.001). However, the IA group had significantly higher PREDICT-PD risk scores than the HC group (median odds 1:103 vs 1:2243, p<0.001) and the PD group had higher risk scores than IA (1:26 vs 1:103, p=0.025).

#### Correlation of duration of anosmia with prodromal features and risk

Shorter duration of anosmia was associated with higher MDS-UPDRS II scores (R^2^=0.15, β=-0.13, p*=*0.024), although there was a significant floor effect (figure 2). This correlation was not affected by including age and gender in the model. Severity on other scales was not associated with reported duration of anosmia.

**Figure 2.**
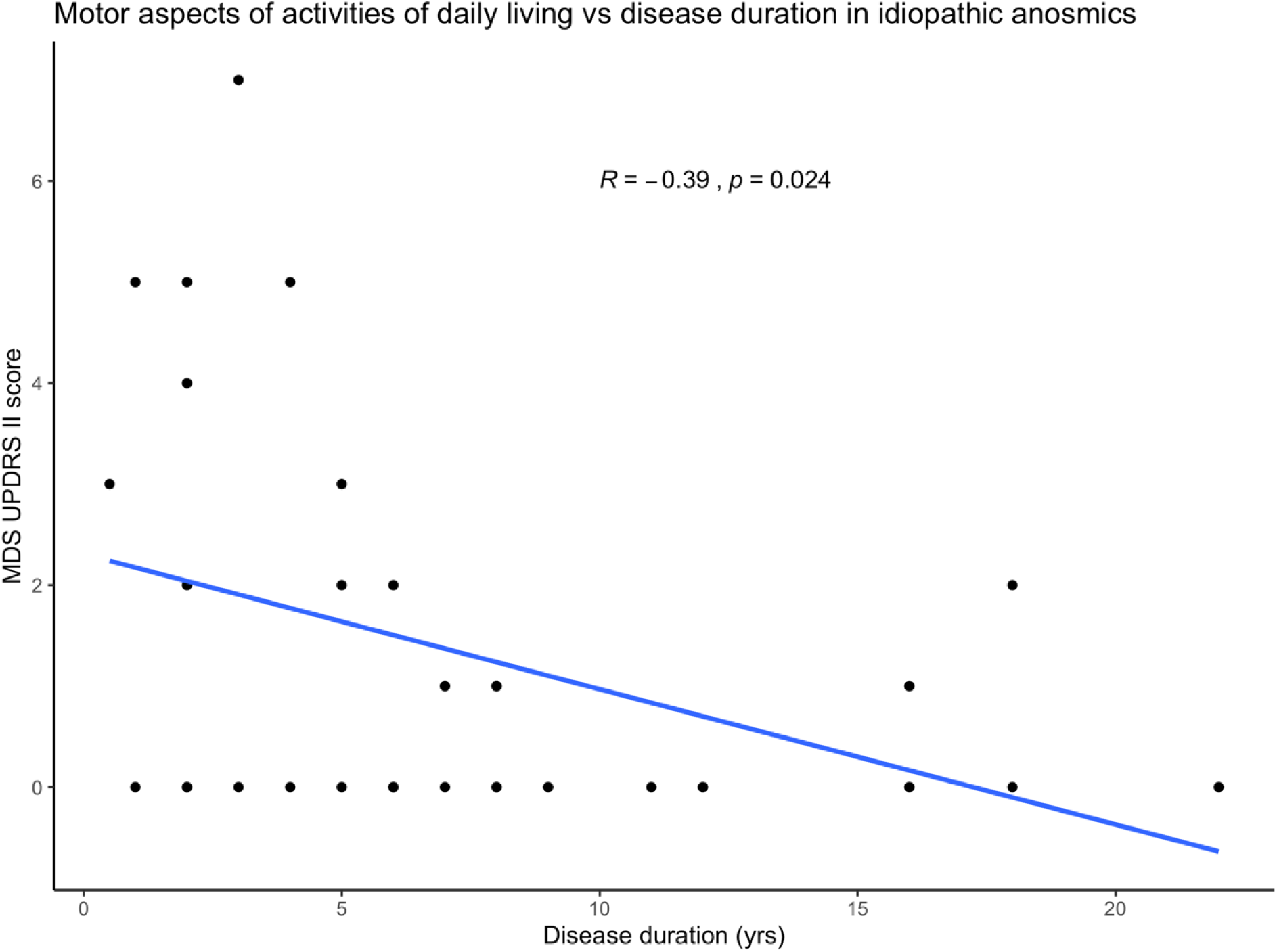
Relationship between idiopathic anosmia disease duration and MDS-UPDRS II (motor aspects of activities of daily living).

## Discussion

### Key results and interpretation

Compared with neurologically healthy older adults, those with IA showed small but significant increases in some non-motor features of PD, including gastrointestinal autonomic symptoms, blood pressure changes, cognition, pain, sleep and depressive symptoms. Trends towards PD were observed across mood, sleep and PREDICT-PD risk scores, which were derived without olfaction data. There was no relationship between duration of anosmia and motor or non-motor features of PD, but shorter duration of anosmia was associated with increased experiences of motor dysfunction, assessed using the MDS-UPDRS II. There was a greater proportion of IA participants who had subthreshold parkinsonism, but this was not statistically significant. Overall, these findings may support the association of IA as a predominantly non-motor prodrome of PD. This is strengthened by follow-up through PREDICT-PD, where 4/42 IA participants have subsequently been diagnosed with PD. The identified associations were relatively subtle, likely reflecting the fact that anosmia can have other causes, even in those where known causes have been excluded. The association of relatively recent anosmia onset with greater motor impairment may suggest that this could be an at-risk group for PD motor symptom onset.

### Cognition

Associations between olfactory dysfunction and cognition have been reported. In a longitudinal study, anosmic PD patients were diagnosed with dementia sooner than hyposmic and normosmic PD patients.^35^ Recently, a cross-sectional analysis looking at olfactory dysfunction and prodromal Lewy body symptoms reported lower median MoCA scores in their IA group than HCs (25 vs 27).^17^ Whilst the present study reported comparable results (IA 25.5 vs HC 27, p=0.019), we did not find that anosmic participants met established cut-offs for MCI or dementia.

### Sleep

The IA group had worse sleep than the HC group. Further, there was no significant difference between the IA and PD patients, suggesting a possible overlap in the sleep dysfunction seen in both groups. There is evidence of olfactory and sleep disturbance co-occurring in other prodromal states of PD. In a prospectively characterised cohort of patients with REM sleep behaviour disorder (RBD), comparing PD converters to non-converters, reduced (and progressively deteriorating) olfaction was present up to a decade prior to conversion.^36^ This was modelled to separate 22 years prior to the diagnosis of PD. However, the neurobiology and neuropathology of RBD likely represents a unique prodromal state, and is a prominent feature of a more malignant phenotype of PD compared to non-RBD PD.^37,38^

### Autonomic

We did not find any relationship between duration of anosmia and global autonomic dysfunction, nor did we find that IA participants had overall more severe dysautonomia than HC. However, within the gastrointestinal subdomain and the SBP change after 3 minutes, there were clinically small but significant differences between IA and HC. Given that the control group are well-matched for age, these GI symptoms may be part of a specific neurodegenerative phenotype rather than ageing related. This subdomain of the SCOPA-AUT includes 3 questions on the oro-pharyngeal aspects of swallowing and hypersalivation, one on early satiety and two on constipation. In a post-hoc analysis of the constipation questions alone, there were low scores overall (median (IQR) IA 0.5 (1), HC 0 (1), PD 1 (1)), with no statistical differences between the groups. This could suggest that the minor differences seen may not be due to Lewy-body pathology in the mesenteric plexus of the lower GI tract but could be related to the more proximal parts of the digestive system, where local spread could be responsible. This requires dedicated clinical and neuropathological investigation in IA and PD patients to explore more thoroughly.

### Motor

We found no differences between IA and HC across the three measures of motor function used (UPDRS-III, writing speed and the TUG). Investigating early motor dysfunction in individuals with RBD, prodromal participants exhibited slower and less rhythmic movements than HC across a battery of motor tasks.^39^ The protocol incorporated two keyboard tapping tasks, the BRAIN test and the Distal Finger Tapping test (DFT) (described elsewhere),^40,41^ which were reported to be more sensitive to subtle motor dysfunction than the UPDRS-III. Although the BRAIN test and DFT now form part of the PREDICT-PD battery, data were not available for participants in the present cohort. Although writing speed measured fine motor control, our motor measures may not have been sensitive enough to capture the earliest changes expected in a prodromal cohort.^42^ Future work should incorporate both the BRAIN test and DFT, alongside TUG, writing speed and UPDRS-III, to capture potential subthreshold motor changes in prodromal groups.

### Pain

The IA group had nominally increased pain compared to HC, similar to the PD group. Pain is a significant symptom in prodromal and early PD, unrelated to motor dysfunction.^43,44^ We found no other studies independently linking anosmia and pain, suggesting this observation may be PD related.

### Depression & Anxiety

No groups showed average scores suggestive of clinical depression or anxiety (i.e. HADS scores >10). However, the IA group had nominally higher depression scores than HC, with no difference to PD participants. A systematic review identified a strong relationship between olfactory dysfunction and depression.^45^ One of the studies reported the prevalence of depression to be between 40% −76% in anosmics with chronic rhinosinusitis, allergic rhinitis or post-viral olfactory loss – although this was a different population to our IA group. This relationship may be multifaceted: loss of enjoyment of food with consequent reduction in socialising around mealtimes, living with a chronic disease and lack of personal hygiene awareness with resultant psychosocial consequences.^46^ Further reduced projections into the limbic and reward pathways may alter higher processing of emotions.^47^ The small differences seen between the IA and HC groups in this study may not represent depression as a prodromal PD symptom, rather a primary relationship between anosmia and depression.

### Relationship with disease duration

We found a significant inverse relationship between motor aspects of activities of daily living (MDS-UPDRS II) and duration of anosmia (R=-0.39, p= 0.020), in keeping with our hypothesis that late onset hyposmia is more likely to be a PD prodrome. Whilst this did not survive multiple comparison correction, the MDS-UPDRS II may not detect changes in early PD,^42^ which would likely be more noticeable in prodromal PD. Smell loss is reported to deteriorate over time after PD diagnosis, in line with disease progression.^48^ These findings, alongside a growing body of literature, suggest that smell loss is a progressive, not static, phenomenon.^35,49,50^ However, there are conflicting data, with several studies showing that smell loss has minimal or no relationship with disease duration.^7,51,52^ An important critique of the Sasaki paper^48^ is that patients in the ‘Early PD’ group contained those who “were not aware…of their parkinsonism, ascribing their symptoms to… ageing… instead”. This arguably describes prodromal PD. Given the smaller proportion of these patients with anosmia, compared to those with more established disease, the findings support the hypothesis that anosmia is a feature of the prodromal phase occurring proximate to PD diagnosis.^42,53^

### Strengths and limitations

A strength is that patients with smell loss had robust assessments excluding other causes of anosmia, leaving a pure ‘idiopathic’ cohort. Whilst such assessments may not be feasible for population-based screening, the results are similar to studies using short smell tests to screen for loss of smell, like PARS.^54^ However, recruitment through subspecialist ENT clinics potentially introduces a recruitment bias into this study e.g. towards those with high motivation and severe symptoms. Masala and colleagues found a strong relationship between apathy and olfactory loss in early PD.^49^ Despite stringent inclusion criteria we may have inadvertently created a selection bias away from those with prodromal PD. We also used a clear definition of IA and controls were selected based on low probability of PD. Nevertheless, despite low risk of PD, some of the HC group may have early neurodegeneration due to other conditions, suggested by some cognitive scores within the MCI range. Another strength is the broad range of assessments across multiple non-motor domains. Several studies examining patients with anosmia have focused purely on motor dysfunction, with or without imaging biomarkers.^11,53,55,56^ Others have included multiple at-risk groups, with no differentiation of syndromes, which potentially conflates prodromal states, with different constellations of symptoms.

The weaknesses of this study include the lack of psychophysical smell testing in all participants. This would have allowed comparison between IA with subthreshold parkinsonism and PD patients with and without olfactory loss, as well as ensuring the control group were not unknowingly anosmic. Quantitative olfactory assessment would have allowed correlation between the degree of olfactory loss and motor and non-motor symptoms within IA. Disease duration of anosmia is prone to recall bias. This is hard to overcome with a cross-sectional study, and the insidious nature of progressive olfactory loss, rather than a ‘time-stamped’ event (such as trauma or infection) to anchor memory of onset. However, the between-group analyses are unlikely to be confounded by this, and although the regression analysis may be affected, the analyses are not likely to be affected by marginal differences, and the trends seen would allow for more error (of some months) in those with longer disease duration who are more prone to recall bias. Longitudinal follow-up would partially mitigate this, highlighting the importance of long-term follow-up to assess conversion to PD. In the PREDICT-PD study, 4/42 of the IA group have developed PD since this study ended, and follow-up of the rest of the group is ongoing. Finally, ascertainment of α-syn SAA status was not possible for the participants in this study. Therefore, it is feasible that there were α-synuclein subgroups within our cohort that may have influenced the associations between prodromal variables and duration of disease, or between groups. The increasing focus on α-synuclein may provide further evidence for the temporal relationship between anosmia, prodromal PD and clinically diagnosed PD, and support the use of disease progression staging frameworks.^60^

## Conclusion

Participants with IA had small but significant differences in cognition, dysautonomic symptoms, pain, depression and PD risk than HC, trended towards worse sleep scores and more closely resembled patients with PD.

Numerically, more IA participants had subthreshold parkinsonism than HC, but total MDS-UPDRS III and functional motor scores were not different. There was a negative relationship between MDS-UPDRS II scores and duration of anosmia.

## Authors’ Roles

Design – RNR, AJN, AS

Execution – RNR, CS, CP, SG, LM

Analysis – RNR, SM, JB, AJN, AS

Writing – RNR, SM

Editing of final version of the manuscript – SM, RNR, CP, CS, JB, AJN, AS

The data that support the findings of this study are available from the corresponding author upon reasonable request.

## Disclosures

The study was funded by Parkinson’s UK grant G1606.

AJN reports grants from Parkinson’s UK, Barts Charity, Cure Parkinson’s, National Institute for Health and Care Research, Innovate UK, Virginia Keiley benefaction, Solvemed, the Medical College of Saint Bartholomew’s Hospital Trust, Alchemab, Aligning Science Across Parkinson’s Global Parkinson’s Genetics Program (ASAP-GP2) and the Michael J Fox Foundation. AJN reports consultancy and personal fees from AstraZeneca, AbbVie, Profile, Roche, Biogen, UCB, Bial, Charco Neurotech, uMedeor, Alchemab, Sosei Heptares and Britannia, outside the submitted work.

AS reports research funding or support from University College London, National Institute of Health (NIHR), National Institute for Health Research ULCH Biomedical Research Centre, the International Parkinson and Movement Disorder Society (IPMDS), the European Commission, Parkinson’s UK, GE Healthcare and the Economic and Social Research Council. AS reports honoraria for consultancy from Biogen, Abbvie, Roche, Bial, GE Healthcare; and license fee payments from the University College London, Royalties from Oxford University Press.

CP is Trustee and the Director of Research and Medical Affairs for Fifth Sense.

SG is a Trustee for the Rhinology and Laryngology Research Fund as well as Abscent. RNR, SM, CS, LM, JB have no relevant disclosures or acknowledgements.

Main body word count: 3688 words (excl. abstract, references, figures and tables)

**Preprint disclaimer:** This preprint reports new research that has not been certified by peer review and should not be used to guide clinical practice.

