## Supplementary materials for "Investigating idiopathic anosmia as a prodromal state of Parkinson’s disease"

**Supplementary Material**

**Table 1.**

| Group | | Normal | Borderline | Abnormal | Median (IQR) |
|---|---|---|---|---|---|
| Depression | HC | 26 | 2 | 0 | 1 (2) |
| | IA | 31 | 2 | 0 | 3 (3) |
| | PD | 9 | 1 | 1 | 6 (4.5) |
| Anxiety | HC | 28 | 0 | 0 | 2.5 (4.25) |
| | IA | 26 | 4 | 3 | 3 (4) |
| | PD | 7 | 3 | 1 | 4 (5.5) |

*Table 1 Ordinal and median scores on the Hospital Anxiety and Depression Scale. Normal <8, Borderline 8-10, Abnormal >10. HC: healthy control, IA: idiopathic anosmia, PD: Parkinson's disease.*

**Table 2.**

| | HC | IA | PD | IA-HC *p* | IA-PD *p* |
|---|---|---|---|---|---|
| Subthreshold parkinsonism (%) | 3 (11.1) | 10 (23.8) | 21 (95.5) | 0.224§ | <0.001§ |
| MDS-UPDRS III | 4 (4) | 3.5 (7) | 31 (16) | 0.852 | <0.001 |
| Writing speed | 1.07 (0.31) | 1.08 (0.36) | 0.77 (0.46) | 0.975 | <0.001 |
| 3m Up & Go | 8.94 (1.24) | 8.93 (1.72) | 10.49 (1.96) | 0.842 | 0.004 |
| MoCA | 27 (3.25) | 25.5 (4) | 26 (3.75) | 0.019 | 0.599 |
| SDMT z-score†* | 0.34 (0.95) | -0.07 (1.1) | -0.04 (0.94) | 0.297 | 0.360 |
| Verbal fluency z-score†* | 0.59 (1.11) | -0.04 (1.11) | 0.83 (2.01) | 0.135 | 0.012 |
| Phonemic fluency z-score†* | 0.28 (1.4) | 0.7 (0.88) | 0.61 (1.19) | 0.092 | 0.771 |
| SCOPA-AUT total† | 7.07 (4.7) | 9.08 (5.04) | 13.2 (6.71) | 0.100 | 0.021 |
| SCOPA-AUT GI | 0 (1.25) | 1 (2) | 4 (4) | 0.035 | <0.001 |
| 1 min SBP pressure change (mmHg) | 3 (13.5) | -2.7 (12.9) | -11.9 (16.2) | 0.090 | 0.035 |
| 3 min SBP change (mmHg) | 5.4 (11.7) | -3.0 (11.5) | -3.7 (18.9) | 0.006 | 0.880 |
| PDSS | 132.5 (20.7) | 124.1 (23.1) | 113.3 (24.5) | 0.015 | 0.084 |
| Pain | 4 (5) | 8 (9) | 7 (9.75) | 0.025 | 0.842 |
| MDS-UPDRS I † | 5.67 (4.69) | 7.29 (4.45) | 11.71 (7.4) | 0.256 | 0.020 |
| MDS-UPDRS II | 0 (1.25) | 1 (2) | 13 (10) | 0.396 | <0.001 |
| HADS depression | 1 (2) | 3 (3) | 6 (4.5) | 0.020 | 0.093 |
| HADS anxiety | 2.5 (4.25) | 3 (4) | 4 (5.5) | 0.093 | 0.663 |
| Risk score | 1:2243 (1832) | 1:103 (171) | 1:26 (54) | <0.001 | 0.025 |
| Natural log risk score | 7.72 (2.46) | 4.63 (2.04) | 3.26 (2.49) | <0.001 | 0.025 |

*Table 2 Comparison of motor and non-motor features between the three groups (All data are median (IQR) with non-parametric comparison, unless indicated by †, where data are mean (SD) with parametric tests); *the higher-level cognitive tasks were started after recruitment with missing data particularly in the anosmic cohort. § - $\chi 2$ test. HC: healthy control, IA: idiopathic anosmia, PD: Parkinson's disease. Some variables had missing data across groups, n per variable and group can be found in supplementary material table 3.*

**Table 3.**

| | HC | IA | PD |
|---|---|---|---|
| Subthreshold parkinsonism | 27 | 42 | 22 |
| MDS-UPDRS III | 28 | 42 | 22 |
| Writing speed | 28 | 39 | 22 |
| 3m Up & Go | 15 | 42 | 18 |
| MoCA | 28 | 42 | 22 |
| SDMT z-score | 11 | 18 | 15 |
| Verbal fluency z-score | 11 | 24 | 18 |
| Phonemic fluency z-score | 24 | 24 | 22 |
| SCOPA-AUT total | 28 | 39 | 22 |
| SCOPA-AUT GI | 28 | 40 | 22 |
| 1 min SBP pressure change | 26 | 39 | 20 |
| 3 min SBP change | 27 | 39 | 20 |
| PDSS | 28 | 39 | 22 |
| Pain | 27 | 42 | 22 |
| MDS-UPDRS I | 15 | 42 | 18 |
| MDS-UPDRS II | 28 | 42 | 22 |
| HADS depression | 28 | 33 | 11 |
| HADS anxiety | 28 | 33 | 11 |
| Risk | 28 | 33 | 11 |

*Table 3. Number of participants contributing data to each variable included in supplementary material table 2. HC: healthy control, IA: idiopathic anosmia, PD: Parkinson's disease. Differences in sample size reflect variable-level missingness across groups.*

The data that support the findings of this study are available from the corresponding author upon reasonable request.
